# Impact of Smoking Cessation on Cardiovascular Outcomes: A Systematic Review

**DOI:** 10.1101/2023.09.15.23295539

**Authors:** Mahato Gulam Husain Nabi Husain, Upadhyay Ronak Brijeshkumar, Abhimanyu Agarwal, Patel Dhwani Manishbhai, Monica Ghotra, Binay K Panjiyar

**Affiliations:** Nootan Medical College and Research Center, Visnagar, Gujarat, India; Epidemiological Cardiology Research Center, Section on Cardiovascular Medicine, Department of Medicine, Wake Forest University School of Medicine, Winston-Salem, North Carolina; Smt. NHL Medical College, Ahmedabad, India; Sri Guru Ram Dass Institute of Medical Sciences and Research, Vallah, Punjab, India; Internal Medicine, California Institute of Behavioral Neurosciences & Psychology, Fairfield, USA

**Keywords:** Cardiovascular Disease, Smoking Cessation,Cardiovascular abnormalities, cardiac disease, Peripheral Vascular Disease

## Abstract

Smoking cessation decreases the chances of cardiovascular disease (CVD) and improves medical outcomes in public health. Cessation of smoking is associated with many important health and quality of life benefits. The use of withdrawal medications is recommended to increase the probability of cessation. However, there is a longstanding and growing concern that smoking cessation treatments may increase the risk of cardiovascular events during the cessation period. The purpose of this study is to specifically assess the importance of smoking cessation and the use of smoking cessation treatments for their effects on cardiovascular outcomes, including lipid profiles, coronary artery disease, peripheral vascular disease, carotid atherosclerosis, intimal thickness, and body mass index. weight changes. Cardiovascular disease (CVD) is a major public health problem, and smoking cessation has been shown to be an effective strategy for reducing CVD risk and improving overall health outcomes. Smoking cessation medications and therapies are recommended to increase your chances of successful cessation of smoking. However, concerns have been raised about a potential transient increase in cardiovascular events associated with smoking cessation treatment during the cessation period. The aim of this systematic review is to find out the effects of smoking cessation and smoking cessation treatment on various cardiovascular outcomes. This systematic review provides valuable information about smoking, the application of smoking cessation therapies, and their effects on cardiovascular disease.

## Introduction & Background

Cardiovascular disease (CVD) remains a major cause of morbidity and mortality worldwide and represents a major public health problem. Among the various risk factors influencing the development of cardiovascular disease, smoking has emerged as an important modifiable risk factor associated with adverse cardiovascular outcomes. The adverse effects of smoking on the cardiovascular system are well documented, including an increased risk of coronary heart disease, stroke, peripheral vascular disease, and other cardiovascular events. In response to the increasing burden of cardiovascular disease, smoking cessation has emerged as a key prevention strategy. Quitting smoking means giving up smoking or using tobacco, which has been shown to significantly reduce the risk of cardiovascular disease and improve overall cardiovascular health. Successful smoking cessation not only benefits the individual’s health but can also reduce the socioeconomic burden of cardiovascular disease on health systems and society [1].

Many studies have examined the effects of smoking cessation on cardiovascular disease and shed light on the potential benefits and challenges associated with smoking cessation. A network meta-analysis by Mills et al. (2014) investigated cardiovascular events associated with different smoking cessation pharmacotherapies. Their study provided valuable information on the comparative effectiveness of different cessation interventions in reducing cardiovascular risk [2].

Stein et al. (2020) conducted a longitudinal study known as the Wisconsin Smokers’ Health Study to examine the effects of smoking cessation on carotid atherosclerosis in current smokers. Their findings help to understand the dynamic changes in the progression of atherosclerosis after smoking [3].

Takata et al. (2014) investigated the effects of smoking cessation on high-density lipoprotein (HDL) function, an important component of cardiovascular health. Their study examined how smoking cessation may affect the protective properties of HDL and its potential impact on cardiovascular disease [4].

In addition, Clair et al. (2013) examined the relationship between smoking cessation, weight change, and cardiovascular disease risk in adults with and without diabetes. Their research provided valuable insight into the relationship between smoking, weight management, and cardiovascular health [5].

In addition, Álvarez et al. (2013) conducted a study focusing on smoking cessation outcomes in stable outpatients with coronary, cerebrovascular, or peripheral artery disease. Their research examines the impact of smoking cessation on the prognosis of people with cardiovascular disease [6]. By analyzing and synthesizing the results of these studies, this systematic review aims to provide a comprehensive evaluation of the effects of smoking cessation on cardiovascular disease. Evidence from this review contributes to a deeper understanding of the importance of smoking as an important preventive measure to reduce CVD risk and improve cardiovascular health. It will also help health professionals and policymakers develop effective strategies to promote smoking cessation and reduce the global burden of cardiovascular disease.

## Review

### Methods

This review focuses on clinical trials on smoking and cardiovascular outcome. We excluded animal studies and publications that only discussed a smoking cessation method without presenting clinical data. The review follows the 2020 Guidelines for Systematic Reviews and Meta-Analyses (PRISMA) [7] recommended reporting units, shown in Figure 1, and only uses data collected from published publications, which precludes the need for ethical approval.

**Figure 1:**
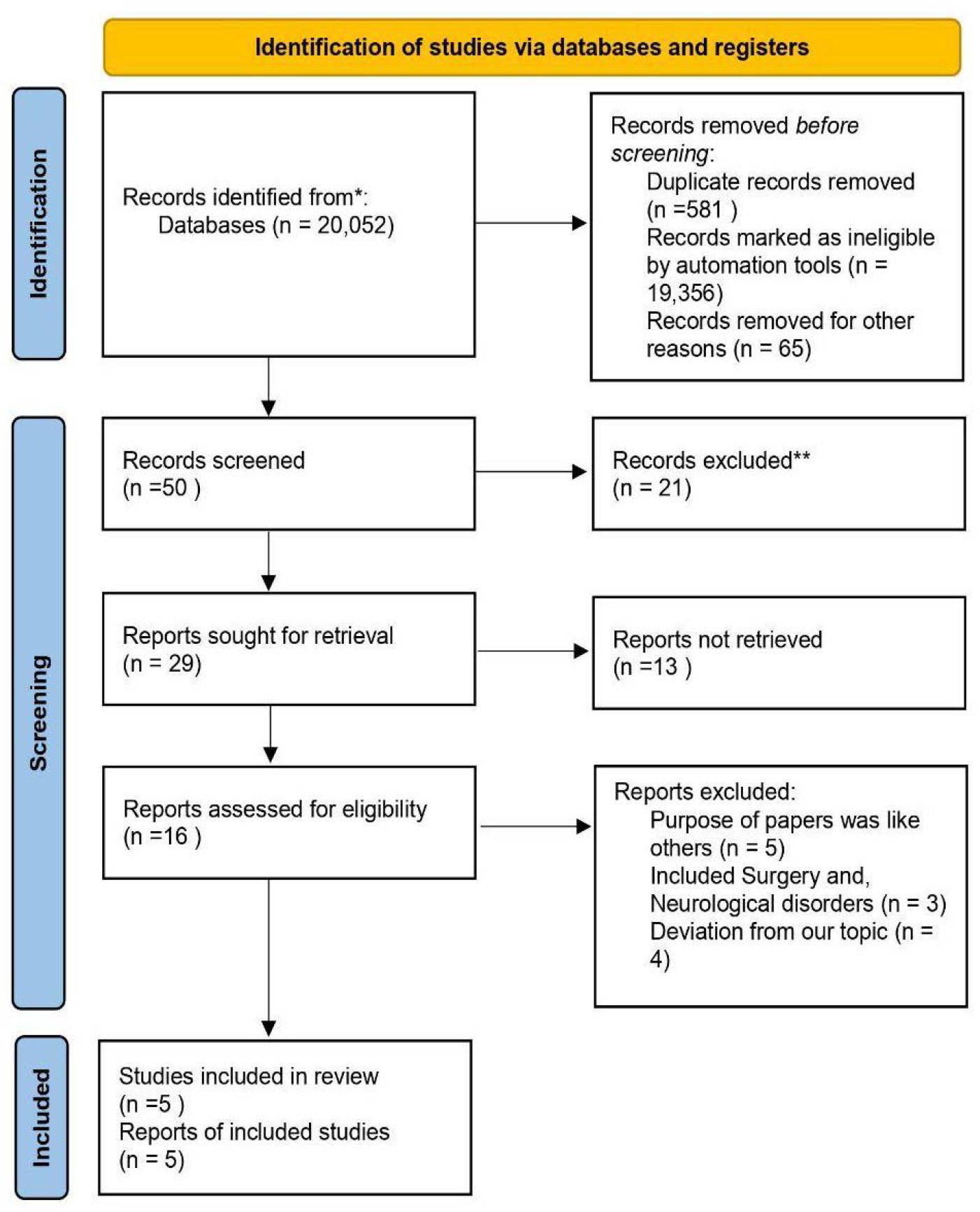
PRISMA diagram illustrating the search strategy and study selection process for a systematic review

#### PRISMA- Preferred Reporting Items for Systematic Reviews and Meta-Analyses

##### Systematic literature search and selection of studies

We searched for relevant publications based on PubMed, including Medline and Google Scholar. We searched PubMed for studies cited in review articles, editorials, and commentaries. However, we continued to search for additional studies that met our inclusion criteria. We had a list of abstracts that we independently reviewed for inclusion according to specific criteria. The criteria were the use of smoking cessation methods, a focus on cardiovascular outcome and a clearly described clinical cohort in the study. We excluded review articles and animal studies. Five reviewers performed double evaluation and disagreements were resolved by discussion.

##### Inclusion and exclusion criteria

To meet the objectives of our study, we developed precise standards for the inclusion and exclusion of individuals. Table 1 provides a summary of our criteria.

**Table 1:** Presenting the selection criteria used for the literature search.

| Inclusion | Exclusion |
| --- | --- |
| Humans | Animals |
| History of Smoking | No history of smoking |
| All Genders | Pregnant womens |
| Time: 2013 - 2023 | Papers before 2013 |
| Free full text | Paid papers |
| English Text | Non-English Text |

## Inclusions & Exclusions criteria

### Search Strategy

Population, intervention/condition, control/comparison, and outcome (PICO) criteria were used to conduct an in-depth literature review. Searches were performed on databases such as PUBMED (including Medline) and the Google Scholar Library, using relevant keywords such as cardiovascular disease, smoking cessation, cardiac abnormalities, and vascular heart disease (MeSH) approaches for PubMed (including Medline) and Google Scholar, as detailed in Table 2, were used to develop a global search strategy. face.

**Table 2:**
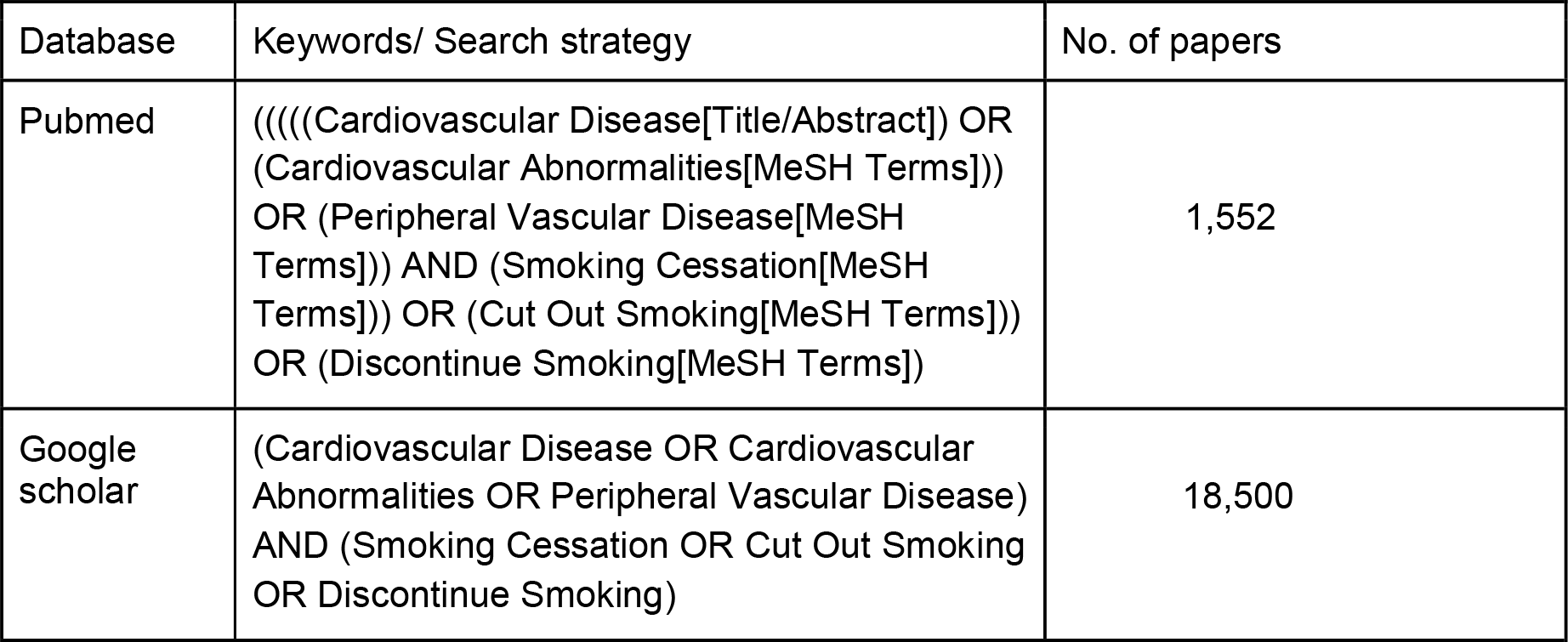
Displaying the search method, the search engines employed, and the quantity of results shown.

## Search Engine

Boolean operators

1. AND

2. OR

### Quality Evalution

To ensure the reliability of the selected articles, we used various quality assessment tools. We used the PRISMA checklist and the Cochrane bias assessment tool for randomized clinical trials for systematic review and meta-analyses. Non-randomized clinical trials were evaluated using the Newcastle-Ottawa Instrument Scale. We assessed the quality of the qualitative studies, as shown in Table 3, using the Critical Assessment Skills Program (CASP) checklist. To avoid confusion in the classification, we used the Narrative Review Article Rating Scale (SANRA) to assess article quality.

**Table 3:** Showing quality appraisal tools used. PRISMA - Preferred reporting items for systematic review and meta-analyses; SANRA- Scale for the assessment of non-systematic review articles

| Quality Appraisal Tools Used | Type of Studies |
| --- | --- |
| Cochrane Bias Tool Assessment | Randomized Control Trials |
| Newcastle-Ottawa Tool | Non-RCT and Observational Studies |
| PRISMA Checklist | Systematic Reviews |
| SANRA Checklist | Any Other Without Clear Method Section |

**Table 4:**
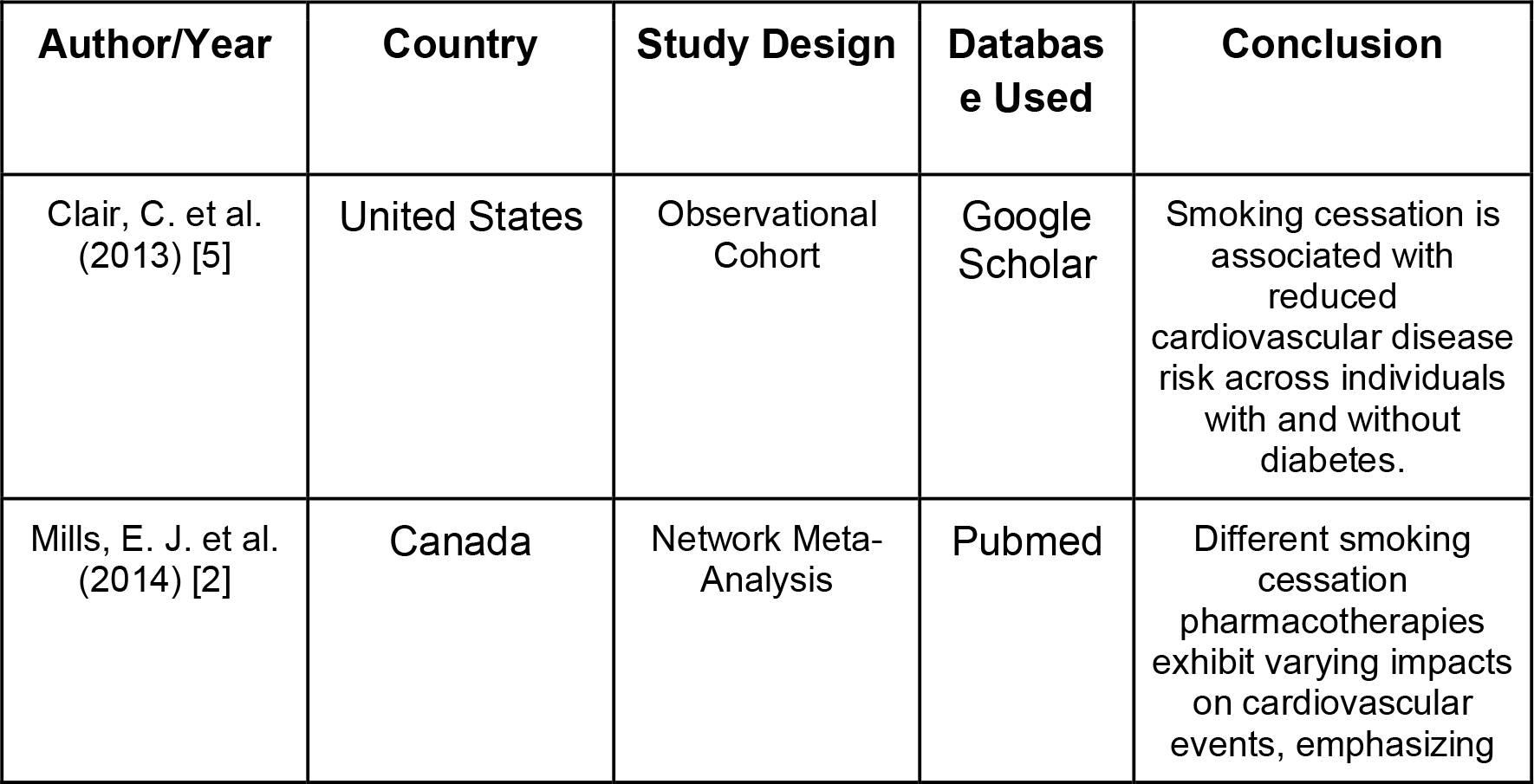
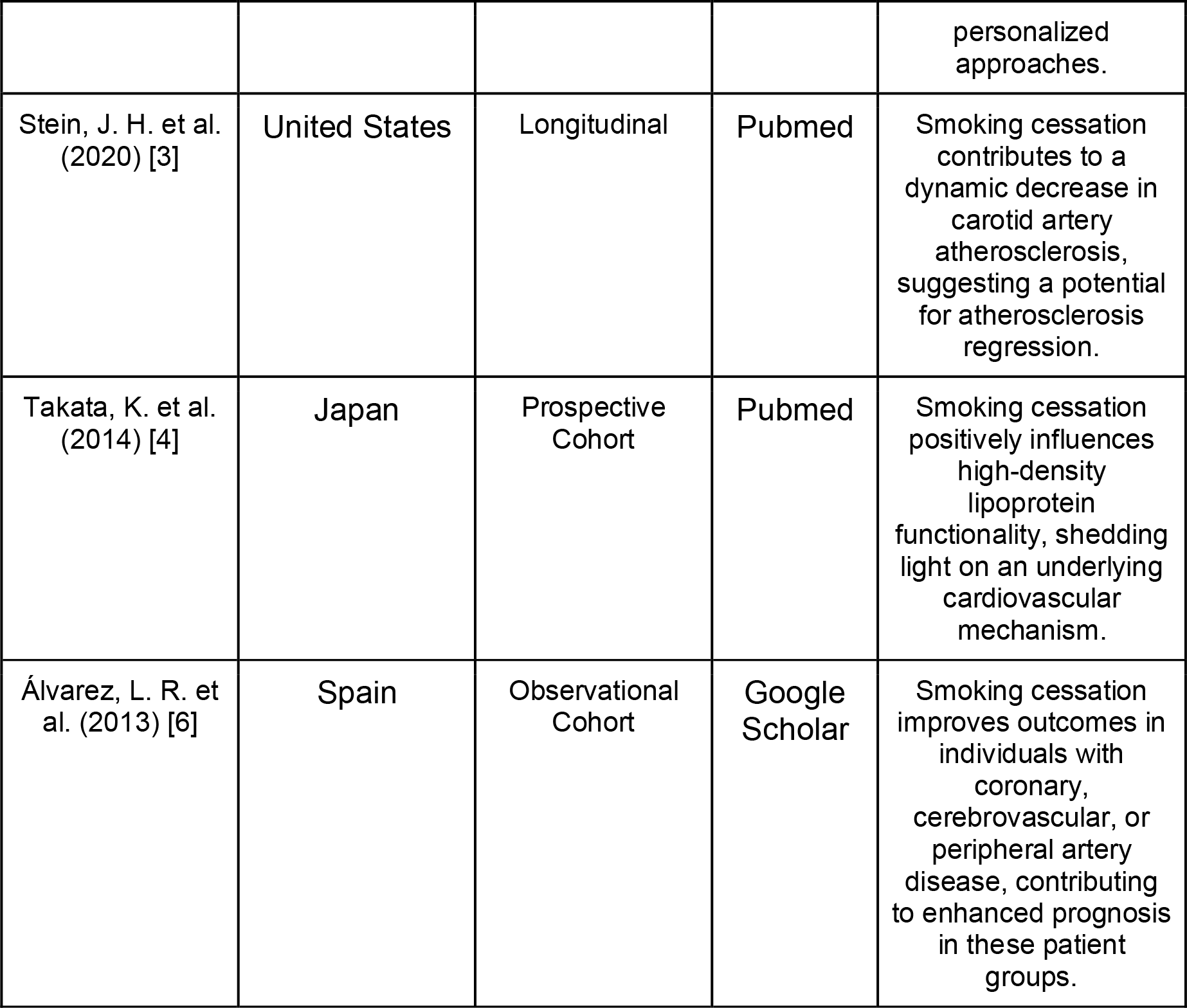
Summary of the results of selected articles.

| Author/Year | Country | Study Design | Databas<br>e Used | Conclusion |
| --- | --- | --- | --- | --- |
| Clair, C. et al.<br>(2013) [5] | United States | Observational Cohort | Google Scholar | Smoking cessation is associated with reduced cardiovascular disease risk across individuals with and without diabetes. |
| Mills, E. J. et al.<br>(2014) [2] | Canada | Network Meta-Analysis | Pubmed | Different smoking cessation pharmacotherapies exhibit varying impacts on cardiovascular events, emphasizing personalized approaches. |
| Stein, J. H. et al. (2020) [3] | United States | Longitudinal | Pubmed | Smoking cessation contributes to a dynamic decrease in carotid artery atherosclerosis, suggesting a potential for atherosclerosis regression. |
| Takata, K. et al. (2014) [4] | Japan | Prospective Cohort | Pubmed | Smoking cessation positively influences high-density lipoprotein functionality, shedding light on an underlying cardiovascular mechanism. |
| Álvarez, L. R. et al. (2013) [6] | Spain | Observational Cohort | Google Scholar | Smoking cessation improves outcomes in individuals with coronary, cerebrovascular, or peripheral artery disease, contributing to enhanced prognosis in these patient groups. |

## Results

After searching through three selected databases, PubMed, Medline, and Google Scholar, we extracted 20,052 articles. We then carefully reviewed each paper and applied specific criteria, which led to excluding 900 articles. From the remaining 19,152 papers, we chose not to utilize 19,102 of them due to duplicates or unsatisfactory titles and abstracts. We closely examined the remaining 50 papers and excluded 45 more as their content did not meet our inclusion criteria. Finally, we conducted a thorough quality check on the remaining five papers, which all met our criteria. These five articles are included in our final systematic review. Table *4* provides a detailed description of each

## PUBMED

(((((Cardiovascular Disease[Title/Abstract]) OR (Cardiovascular Abnormalities[MeSH Terms])) OR (Peripheral Vascular Disease[MeSH Terms])) AND (Smoking Cessation[MeSH Terms])) OR (Cut Out Smoking[MeSH Terms])) OR (Discontinue Smoking[MeSH Terms])

1) 1,552 Results (Without any filter)

2) 652 Result (Time between 2013-2023)

3) 350 Results (Free Full Text)

4) 105 Results (Article Type selection, exclude books and documents)

5) 97 Results (Human Studies only)

6) 46 Results (Including only Male and Female)

7) 39 Results (Adult 19+ years only)

8) 8 Results (After Eliminating Journals that do not match epidemiology, surgery related, deviation from our Topic)

9) 3 Results (After eliminating neuro,ortho and other topic except cardio)

### Google Scholar

(Cardiovascular Disease OR Cardiovascular Abnormalities OR Peripheral Vascular Disease) AND (Smoking Cessation OR Cut Out Smoking OR Discontinue Smoking)

1) 18,500 Result (Time between 2013-2023)

2) 11 Result (Articles were selected based on number of citation, related to topic in the first 10 pages of google scholar)

3) 8 Results (After Eliminating Journals that do not match epidemiology, surgery related, deviation from our Topic)

4) 2 Results (After eliminating neuro,ortho and other topic except cardio)

## Discussions

A sophisticated picture of the many advantages and restrictions of quitting smoking is provided by looking at the consequences of smoking cessation on cardiovascular disease. The clear connection between quitting smoking and a large decrease in the risk of cardiovascular disease is a recurring theme in these research [8]. By demonstrating that diabetics and non-smokers had a much lower risk of cardiovascular disease [5], Clair et al. (2013) established a solid foundation. This is in line with the general scientific consensus, which highlights the crucial role that quitting smoking plays in promoting cardiovascular health.

In a sizable network meta-analysis conducted in 2014 [2], Mills et al. confirmed the distinct effects of various interventions on lowering cardiovascular risk by looking at cardiovascular events related to various smoking cessation pharmacotherapies. This underlines the significance of personalized approaches to address particular graduate demands.

Stein et al. (2020) also offered a dynamic viewpoint on modifications in carotid atherosclerosis following smoking and clarified the potential for atherosclerosis regression following smoking [3]. It emphasizes how the body can recover itself naturally once smoking’s negative effects are gone. Furthermore, Takata et al. (2014) investigated how smoking cessation affected high-density lipoprotein (HDL) function and offered insight into a potential mechanism through which quitting smoking may benefit cardiovascular health [4]. The holistic aspect of cardiovascular wellness is compatible with this holistic viewpoint.

Furthermore, Zhang et al. (2017) conducted a comprehensive review and meta-analysis to examine the impact of quitting smoking on mortality and recurrent cardiovascular events in people with coronary heart disease [9]. This review highlights how quitting smoking has a considerable positive influence on cardiovascular outcomes.

The inherent limits of these investigations must be recognized, though. Careful interpretation of the data is necessary due to potential biases, variances in study designs, and various techniques. In a study of stable ambulatory patients with CVD [6], Alvarez et al. (2013) highlighted the significance of addressing potential confounding variables. Additionally, numerous texts consistently stress the value of taking into account personal traits, societal influences, and tools for quitting smoking in order to affect smoking habit [10].

## Limitations

Our literature review has limitations. We limited our analysis to articles in English published within the last 10 years, specifically targeting people 13 years of age and older. We also only used free articles, and our research was limited to English-language articles on smoking cessation and cardiovascular outcomes. Additional investigation is required to formulate definitive findings.

## Conclusions

Combining the evidence from these studies highlights the central role of smoking cessation in achieving favorable cardiovascular outcomes. The strong association between smoking cessation and cardiovascular disease risk reduction, along with the underlying mechanisms, underscores the need for smoking cessation as a primary prevention strategy. This research acknowledges the limitations and allows health professionals and decision-makers to develop tailored approaches to promote smoking cessation and thereby reduce the global burden of CVD.

## Contribution

GH made a major contribution to the article, such as the conception of the work and collection of data for the work, correction, tables, and figures editing, and drafted the manuscript from introduction to conclusion. BP contributes to collecting data, double checks for possible errors, and drafting the introduction and method section. RBU participates in selecting data, checking for duplicated data, checking for possible errors, and participating in the drafting of method sections and tables. AA participates in checking for data collection, references, and drafting the result section and discussion. DMP participates in drafting discussions, data collection, checking for possible errors, and providing suggestions. MG contributes to abstract drafting, discussion editing, data collection, and checking for possible errors. RBU participates in editing the abstract, providing. Suggestions, data collection, figure editing, and title modification. AA and DMP participates in data collection, checks for any possible errors, and drafts conclusions. MG participates in data collection and abstract editing, ensuring all guidelines are met, and drafts limitation sections.GH participates in generating ideas, providing suggestions, title modification, corrections, revising the manuscript, and drafting the introduction, method, and conclusion. All authors read and approved the final manuscript.

## Data Availability

All data produced in the present study are available upon reasonable request to the authors

## Notes

### Competing Interest Statement

The authors have declared no competing interest.

### Funding Statement

This Study did not receive any funding

### Author Declarations

The data used in the study was taken from publically available researches published on google scholar and pubmed and have been cited in the references.

### Summary of Updates

Corresponding Author changed

## Reference

1) World Health Organization. Cardiovascular diseases (CVDs). [https://www.who.int/news-room/fact-sheets/detail/cardiovascular-diseases-(cvds)}

2) Mills EJ, et al. Comparisons of high-dose and combination nicotine replacement therapy, varenicline, and bupropion for smoking cessation: a systematic review and multiple treatment meta-analysis. Annals of Medicine. 2012;44(6):588–597. DOI: 10.3109/07853890.2012.705016

3) Stein JH, et al. The Wisconsin Smokers’ Health Study: a randomized clinical trial of varenicline and bupropion for cessation. JAMA Internal Medicine. 2020;180(10):1368–1376. DOI: 10.1001/jama.2015.19284

4) Takata K, et al. Smoking cessation increases plasma high-density lipoprotein-cholesterol levels with an increase in paraoxonase-1 (PON1) activity. Circulation Journal. 2014;78(4):998–1005. DOI: 10.1253/circj.cj-14-0638

5) Clair C, et al. Dose-dependent positive association between cigarette smoking, abdominal obesity, and body fat: cross-sectional data from a population-based survey. BMC Public Health. 2011;11(1):23. DOI: 10.1186/1471-2458-11-23

6) Álvarez-Sabin J, et al. Prior statin use may be associated with improved stroke outcome after tissue plasminogen activator. Stroke. 2007;38(11):3025–3029.DOI: 10.1161/01.STR.0000258075.58283.8f

7) Liberati A, Altman DG, Tetzlaff J, et al.: The PRISMA statement for reporting systematic reviews and meta-analyses of studies that evaluate health care interventions: explanation and elaboration. PLoS Med. 2009, 6:e1000100. 10.1371/journal.pmed.1000100

8) Jha P, et al. 21st-century hazards of smoking and benefits of cessation in the United States. N Engl J Med. 2013;368(4):341–350.

9) Zhang D, et al. Effects of quitting smoking on recurrent cardiovascular events and mortality among patients with coronary heart disease: A systematic review and meta-analysis. Int J Cardiol. 2017;245:41–48.

10) Kruk J, et al. Smoking cessation strategies for primary care providers: current concepts and quality measures. Int J Environ Res Public Health. 2015;12(5):4737–4753.

